# Pixel-Based Skin Tone Estimation on Dermoscopy: A Dual-Rater MST Benchmark and Feasibility Study

**DOI:** 10.64898/2026.05.13.26353004

**Authors:** Amindu Kumarasinghe, Vinh Bui, Reza Ghanbarzadeh

## Abstract

Skin-tone labels are absent from public dermoscopy benchmarks such as the International Skin Imaging Collaboration (ISIC), making it impossible to audit whether clinical AI performs equitably across skin tones. While several recent works estimate skin tone automatically from clinical photography and selfies, we ask whether this approach is feasible on dermoscopy, the primary imaging modality of these benchmarks.

To answer this, we make three main contributions. First, we release MST-Derm, a dual-rater Monk Skin Tone (MST) annotation benchmark on 500 ISIC 2018 images. Raters were given an explicit *unrateable* option for crops where the skin surrounding the lesion was too occluded to label confidently. We find that 60% of images were marked unrateable, yielding a 193-image consensus subset (quadratic-weighted Cohen’s Kappa = 0.82).

Second, we conduct a systematic feasibility study of three pixel-based MST annotation pipelines spanning the principal families in prior work: palette matching in perceptual colour space, robust colour statistics, and projection to a 1D colorimetric scalar. All three pipelines produce ordinal signal above chance (95% confidence intervals on quadratic-weighted Kappa exclude zero). However, ISIC 2018’s extreme light-skin bias leaves 82% of the evaluation set at MST 2, giving a constant “always predict MST 2” baseline an accuracy floor the methods cannot overcome. To separate algorithmic signal from dataset bias, we evaluate on a class-balanced subset. The best method reaches quadratic-weighted Kappa = 0.43 against the trivial baseline of Kappa = 0.00, confirming the signal is genuine.

Third, we diagnose this performance ceiling. We trace the bottleneck to two causes: dermoscopy’s specialised illumination physically compresses the colour range on which lighter skin tones differ, and ISIC’s dataset skew makes standard absolute-accuracy metrics uninformative.

We conclude that while pixel-based colour features carry real MST signal on dermoscopy, current performance is insufficient for autonomous annotation. We release the benchmark, annotation protocol, all prediction runs, and analysis code to facilitate the development of robust skin-tone estimators, a vital prerequisite for accurately auditing fairness and mitigating bias in dermatological machine learning.

## 1 Introduction

Public dermatological image datasets underpin a growing body of clinical machine learning, and the skin-tone distribution of those datasets is a foundational variable for fairness audits, training stratification, and the evaluation of synthetic skin-lesion generators [6, 8]. The Monk Skin Tone (MST) scale [11, 12] has emerged as an increasingly adopted reporting standard in this space. Existing skin-tone estimators, including the individual typology angle (ITA) on *L*\*, *b*\* [3, 9], supervised classifiers trained on annotated face or full-body images [6], and palette-matching approaches in CIE Lab [15], were developed primarily on clinical photography and selfies. Whether they transfer to dermoscopy is, to our knowledge, untested in the open literature.

Dermoscopy is a very different visual modality. It is captured under cross-polarized LED illumination through immersion fluid or a contact plate, which suppresses surface glare and emphasises sub-surface structure [1]. The same illumination desaturates the chromatic axes *a*\* and *b*\* on which lighter skin tones are already difficult to distinguish. Beyond the modality shift, public dermoscopy benchmarks such as ISIC 2018 [4] draw primarily from lighter-skinned populations [6], and dermoscopy crops a small field of view around the lesion, so the visible perilesional (surrounding-lesion) skin available for colour estimation is small and frequently hair- or ink-occluded.

To determine whether pixel-based annotation pipelines can overcome these optical and distributional constraints, we make three contributions:

1. **MST-Derm: a dual-rater MST annotation benchmark on 500 ISIC 2018 dermoscopy images**. Each image is labelled independently by two raters following a written protocol, with an explicit *unrateable* (NA) option for crops where the visible perilesional skin is too occluded to commit to a swatch. Inter-rater reliability on the 193 images where both raters assigned a numeric label (the remainder were marked unrateable) is quadratic-weighted Cohen’s kappa (*κ* = 0.82), in the same range as published MST inter-rater reliability on still photography (ICC = 0.86–0.94; 12).
2. **A feasibility study across three methodological families**. We implement three pixel-based MST annotation pipelines sharing a common front-end (lesion segmentation, perilesional skin sampling, and skin filtering) and a domain calibration step that replaces canonical MST reference colours with anchors extracted from the dermoscopy images themselves. The three pipelines span the principal families in prior work: palette matching in perceptual colour space (Palette), robust colour statistics (TrimLab), and projection to a 1D colorimetric scalar (ITA). All three produce statistically significant ordinal signal on the 170-image evaluation set (95% CI on QWK: Palette [0.07, 0.30], TrimLab [0.09, 0.34], ITA [0.07, 0.33]).On a class-balanced evaluation (equal resampling from each MST class to remove the dataset skew), the best method reaches quadratic-weighted *κ* = 0.43 against a trivial baseline of *κ* = 0.00. On the full evaluation set (82% MST 2), however, all three are dominated by a constant “always predict MST 2” baseline on exact-match and error metrics, because the class skew gives any constant predictor an accuracy floor the methods cannot overcome.
3. **A diagnosis of the performance bottleneck**. The gap between real signal and sufficient performance has two causes. First, dermoscopy’s specialised illumination compresses the colour range on which lighter skin tones differ, causing the extracted colours for MST 1, 2, and 3 to overlap heavily regardless of which pipeline is used (Section 4.3). Second, ISIC’s light-skin bias, compounded by the high rate of unrateable images on darker and hair-dense crops, leaves 82% of the evaluation set at MST 2 and no images above MST 5. The class-balanced evaluation (Section 4.2) shows that removing this skew raises the best method’s *κ* to 0.43 against a trivial baseline of zero, establishing dataset representativeness as the primary bottleneck.

The feasibility finding is specific to dermoscopy on ISIC and to pixel-based methods. We do not claim that no method can label dermoscopy images by skin tone, only that three substantially different pixel-based methods, one per principal family in the prior literature, produce real but insufficient signal on this modality at the current dataset scale. We discuss what this implies for other modalities, where the illumination-induced colour compression is absent and where pixel-based MST estimation is likely still viable, in Section 5.

The remainder of the paper is organised as follows. Section 2 reviews the MST scale, CIE Lab, dermoscopy as an imaging modality, and the three families of prior automated skin-tone estimators. Section 3 describes the MST-Derm benchmark, its annotation protocol, and inter-rater reliability. Section 4 specifies the three pixel-based pipelines and reports their performance against constant and random-guess baselines. Section 5 diagnoses the performance bottleneck, scopes the feasibility finding, and identifies future directions. Section 6 concludes.

## 2 Background

This section reviews the four ingredients the feasibility study depends on: the Monk Skin Tone scale, the CIE Lab colour space and CIEDE2000 distance, the optical properties of dermoscopy that distinguish it from clinical photography, and prior automated skin-tone annotation work.

### 2.1 The Monk Skin Tone scale

The Monk Skin Tone (MST) scale [12] is a ten-category ordinal palette designed to cover the perceptual range of human skin more uniformly than the six-category Fitzpatrick scale [7], which was originally developed to predict UV response rather than to describe skin colour. Each MST category is anchored by an sRGB reference colour (Figure 1). The original scale validation reports human inter-rater reliability of ICC = 0.86–0.94 across annotation conditions on still photography [12]; we use this as an order-of-magnitude reference when reporting our own rater agreement (measured as quadratic-weighted Cohen’s *κ*) on dermoscopy in Section 3.4. Use of MST in dermatology specifically has been advocated but is under-validated [11]. We treat MST as a label space, not a biological ground truth: two raters shown the same patch can legitimately disagree by one shade, and our evaluation reflects this by reporting within-one-shade agreement alongside exact match.

**Figure 1:**
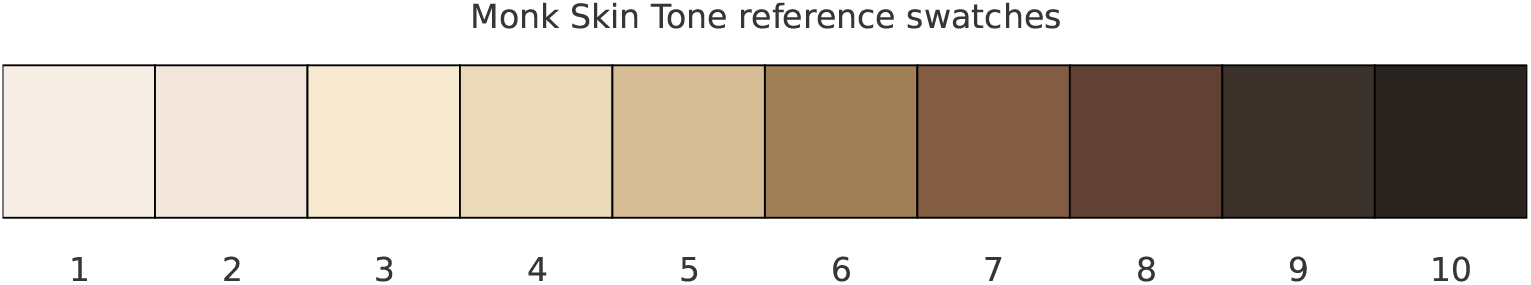
Monk Skin Tone reference swatches. The ten sRGB anchor colours defining the MST scale, ordered from lightest (MST 1) to darkest (MST 10). Pixel-based MST annotation methods typically extract a dominant skin colour from an input image and label it with the index of the nearest anchor under a perceptual distance.

### 2.2 CIE Lab and CIEDE2000

CIE Lab [5] is a perceptually uniform colour space: unlike sRGB, equal Euclidean distances in Lab correspond approximately to equal perceived colour differences. It separates lightness (*L*\*) from two chromatic axes (*a*\*, *b*\*), and CIEDE2000 [10, 13] corrects for residual nonlinearities in Lab, particularly in the blue region and at low chroma. Converting the canonical sRGB anchors [12] to CIE Lab reveals that MST 1–3 are separated primarily along the *b*\* axis: their *L*\* values span about 2 units (from 92.3 to 94.2), *a*\* spans about 2 units (from 0.2 to 2.1), but *b*\* ranges from 5.4 (MST 1) to 14.2 (MST 3). The Euclidean Lab distance between the MST 1 and MST 2 anchors is only 2.7 units, smaller than the typical within-image *a*\*/*b*\* extraction noise on dermoscopy that we report below. Any pipeline that estimates MST in CIE Lab is therefore critically dependent on the integrity of *a*\* and *b*\* in the input image.

### 2.3 Dermoscopy as an imaging modality

Dermoscopy is captured at close range through a contact plate or immersion fluid under cross-polarized LED illumination. The cross-polarization filter suppresses surface specular reflection, exposing sub-surface melanin and vascular structure [1]. The resulting images have three properties relevant to skin-tone estimation:

1. **Compressed chromatic dynamic range**. Suppression of surface specular reflectance is expected to reduce skin surface chromaticity [1]. Our results (Section 4.3) are consistent with this: the extracted *b*\* values for MST 1–3 overlap heavily, collapsing the separation that exists in the MST reference colours.
2. **Small skin-to-lesion ratio**. Dermoscopes crop tightly around the lesion. The visible perilesional skin is typically a thin ring, often partly occluded by hair or ink markings, and shrinks with lesion size.
3. **Modality-specific artifacts**. Hair, ruler markings, and surgical markings [6] contaminate the perilesional ring. These artifacts must be filtered out to isolate the non-diseased skin before objective colour estimation can be performed [9].

These properties are specific to contact dermoscopy and absent from the selfies, stock photography, and clinical images on which prior pixel-based skin-tone work has been conducted [6, 9, 15].

### 2.4 Lesion segmentation

Lesion segmentation in dermoscopy is a mature task with strong public baselines [4]. We use DermoSegDiff [2], a diffusion-based segmentation model trained on the ISIC 2018 lesion segmentation challenge [4], which reports a Dice coefficient of 0.9005 on the ISIC 2018 test set

(five-sample ensemble, DermoSegDiff-A variant), where Dice measures mask overlap on a 0–1 scale. Segmentation quality is not the performance bottleneck; we discuss this in Section 5.

### 2.5 Automated skin-tone annotation: prior approaches

Prior automated approaches to dermatological skin-tone annotation fall into three groups.

#### Individual typology angle (ITA)

ITA [3, 9] is a continuous scalar derived from *L*\* and *b*\*, defined as ITA = arctan((*L*\* −50)/b*)·180*/π*. It is simple, requires no training, and has been used to bin subjects into five ITA-based categories [9]. ITA inherits the same dependence on *b*\* that we identify as a source of chromatic compression in dermoscopy, and we expect it to face the same performance ceiling as palette matching on this modality.

#### Supervised classifiers

CNNs fine-tuned on Fitzpatrick-labelled image datasets can classify skin tone directly from pixels [14]. This approach is accurate within its training modality, but transfer to other modalities is not guaranteed; it requires per-modality labelled training data, which does not exist for dermoscopy MST annotation.

#### Palette matching

Thong et al. [15] extract a representative skin colour and map it to a continuous, multidimensional colorimetric space (using lightness and hue). This is the direct ancestor of the pipeline we evaluate. Their evaluation is on selfies and stock photography, transfer to dermoscopy was not tested.

To our knowledge, no prior published work evaluates pixel-based MST estimation specifically on dermoscopy with dual-rater human ground truth. We address this gap.

## 3 The MST-Derm Benchmark

This section describes the dataset, the dual-rater annotation protocol, the inter-rater reliability we observe, and the label distribution. The benchmark is the paper’s primary contribution and defines the evaluation set used in Section 4.

### 3.1 Image source and sampling

We sampled 500 dermoscopy images uniformly at random, without replacement, from the public ISIC 2018 skin lesion classification challenge training set [4], which contains 2,594 images. We did not condition the sample on skin tone. The sample therefore inherits ISIC 2018’s underlying skin-tone distribution, which prior work has shown is heavily biased toward lighter skin [6, 8].

### 3.2 Annotation protocol

Two raters independently labelled all 500 images. Both were non-clinicians; their demographics, including self-reported skin tone, were not recorded. Raters followed the same written protocol (released with the data), which fixes the MST reference swatches as displayed colour patches at a calibrated screen brightness and admits an explicit *NA* (*unrateable*) label for images where the visible perilesional skin is too occluded to support a confident swatch match. The full protocol text is reproduced in Section A; we discuss the implications of rater pool composition in Section 5.4.

#### The NA convention

The NA option is invoked when the visible perilesional skin is too occluded to support a confident swatch match. Occlusion sources include dense terminal hair covering the perilesional ring, surgical ink markings, ruler-mark overlays, immersion-fluid bubbles, and aggressive cropping that leaves no perilesional skin in frame. Colour-contamination sources that also trigger NA include peri-lesional inflammation that tints the visible skin. The protocol does not penalise NA usage: raters were instructed that abstaining from a guess is preferable to producing a low-confidence numeric label, because downstream uses (subgroup audits, dataset stratification) treat numeric labels as ground truth.

Raters viewed the full dermoscopy image at full screen and could zoom freely. They were not shown the lesion mask or the perilesional ring used by the algorithm.

### 3.3 Label distribution and the high NA rate

Rater H1 labelled 197/500 images with a numeric MST value and 303/500 (60.6%) as NA; rater H2 labelled 205/500 numerically and 295/500 (59.0%) as NA. The dual-rater consensus subset (images where both raters produced a numeric label) contains 193 images. The per-rater distribution is shown in Figure 2.

**Figure 2:**
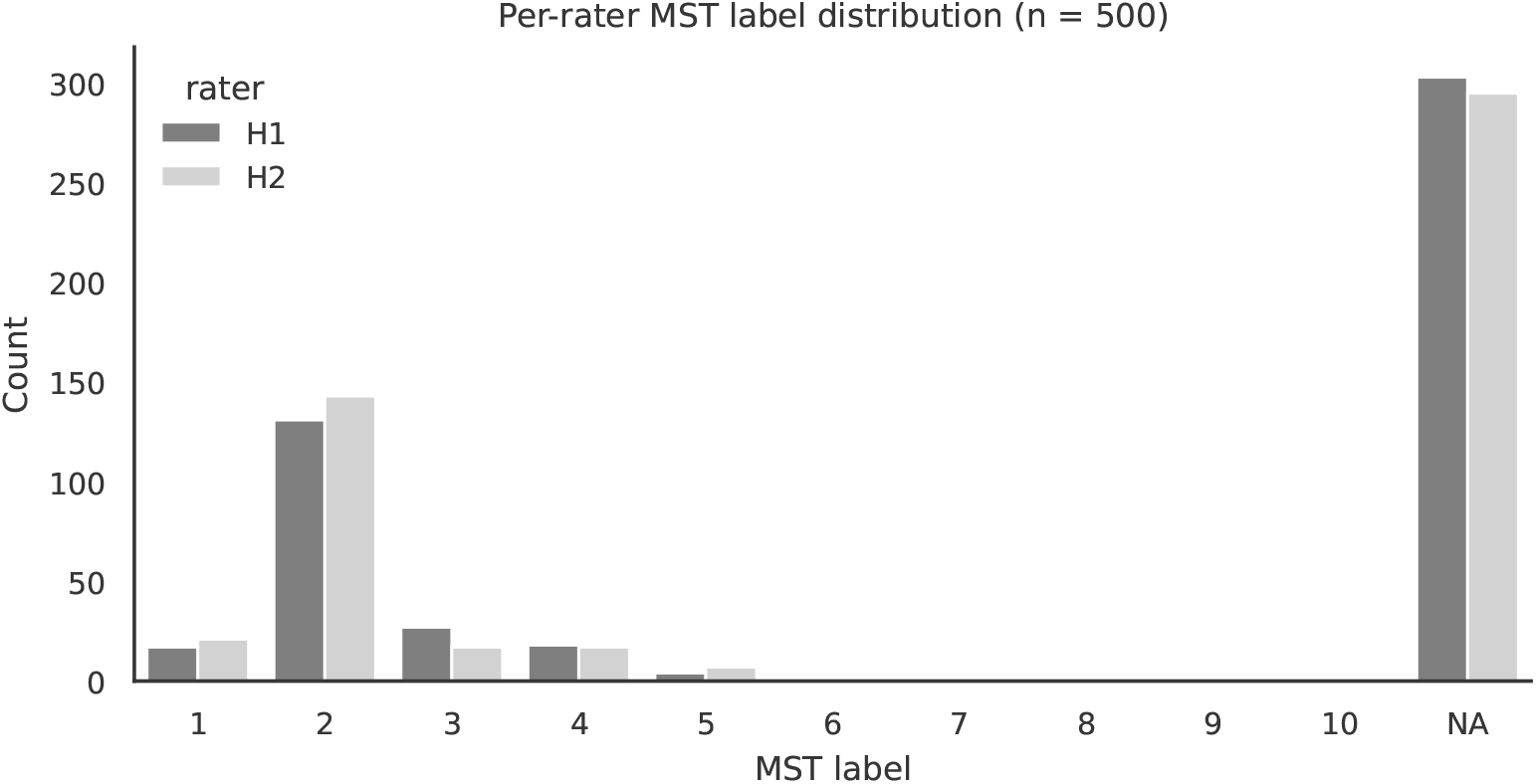
Per-rater MST label distribution across the 500-image benchmark. Both raters mark the majority of images as *unrateable* (NA), reflecting how much of the perilesional skin in ISIC 2018 crops is occluded by hair, ink, or aggressive cropping. Among numerically rated images, mass concentrates on MST 2; no image in the dual-rater consensus subset received a label above MST 5.

The 60% NA rate is itself a finding. It implies that on a majority of ISIC 2018 dermoscopy crops there is insufficient visible perilesional skin for a human, free to zoom and hold the swatch reference adjacent to the image, to commit to a numeric MST label. Any automated method that produces a numeric label for every image is, by construction, producing labels for images that humans found unrateable. This study does not evaluate algorithm output on the NA partition, because there is no human ground truth to evaluate against. We discuss the consequences of this asymmetry for downstream applications in Section 5.2.

The label distribution on the consensus subset is highly skewed: 14 images at MST 1, 144 at MST 2, 14 at MST 3, 18 at MST 4, 3 at MST 5, and 0 at MST 6–10. This is consistent with prior reports that ISIC underrepresents darker skin tones [6], compounded by the NA rule removing the most occluded crops from the rateable set. We treat this skew not as a flaw of the protocol but as a property of the underlying dataset that the benchmark faithfully exposes. It has direct consequences for the main result we develop in Section 4.2.

### 3.4 Inter-rater reliability

Table 1 reports inter-rater reliability on the 193-image consensus subset. The two raters agree exactly on 86% of images (hereafter the *exact-agreement subset*), agree within one shade on 97.4%, and reach a quadratic-weighted Cohen’s *κ* = 0.82. This is in the same range as the ICC = 0.86–0.94 reported for MST on still photography [12] (the metrics differ, the comparison is order-of-magnitude only) and confirms that the labelling task on dermoscopy, when raters are free to abstain, is well-defined enough to support a reproducible benchmark.

**Table 1:**
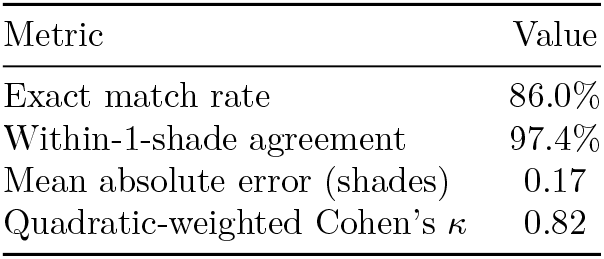
Human inter-rater reliability on the 193-image consensus subset. Quadratic-weighted Cohen’s *κ* penalises large disagreements more than small ones. *N*=193.

| Metric | Value |
| --- | --- |
| Exact match rate | 86.0% |
| Within-1-shade agreement | 97.4% |
| Mean absolute error (shades) | 0.17 |
| Quadratic-weighted Cohen’s $\kappa$ | 0.82 |

Figure 3 shows the inter-rater confusion matrix restricted to MST 1–5 (the only categories that appear in the consensus subset). Rater disagreements are concentrated on MST 2/3 and MST 3/4, consistent with the small inter-anchor distance at the lighter end of the scale.

**Figure 3:**
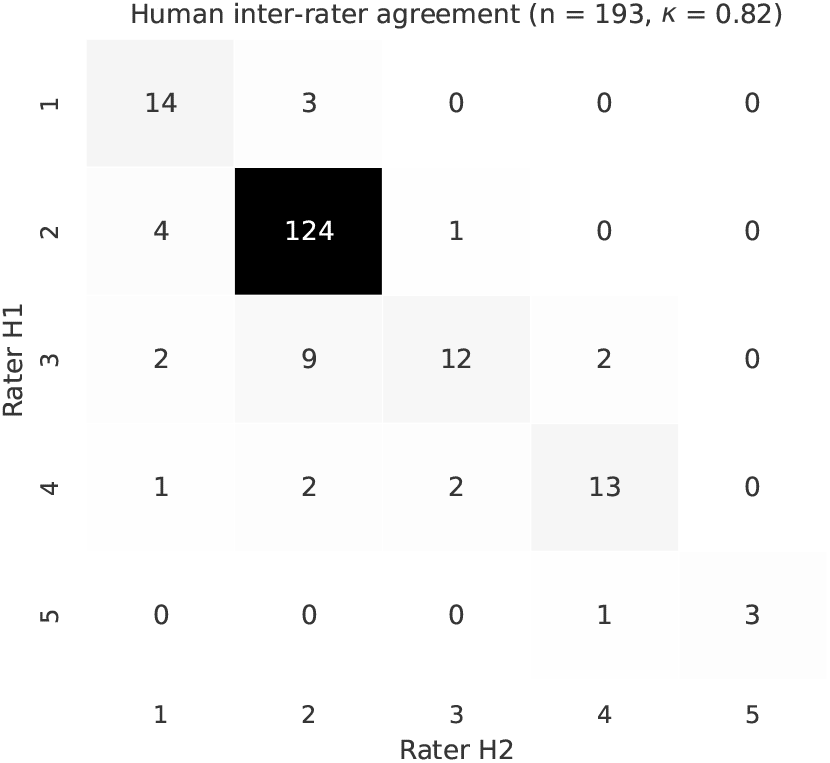
Human inter-rater confusion matrix. Counts of image pairs by rater H1 (rows) and rater H2 (columns) on the 193-image consensus subset, restricted to MST 1–5 (the only categories that appear). Disagreements concentrate on adjacent shades.

### 3.5 Calibration–evaluation split

The pipeline we evaluate in Section 4 includes an optional calibration step that re-estimates MST anchor colours from a small set of archetype images per category. To avoid evaluating on calibration data, we draw a stratified random sample of archetypes from the exact-agreement subset (images where the two raters’ numeric labels match) and hold out the remaining 170 of the 193 consensus images as the evaluation set. The selection is reproducible from a fixed seed; the exact split, the per-class archetype lists, and the eval set are released as `data/archetype_selection/`. The calibration archetypes total 23 images, drawn from MST 1–5; the evaluation set therefore contains 170 images with the consensus distribution shown in Table 2.

**Table 2:** Consensus class distribution on the 170-image evaluation set. The evaluation set is held out from the 23 calibration archetypes. No images above MST 5 appear in the consensus subset and therefore none in the evaluation set.

| Class | 1 | 2 | 3 | 4 | 5 | 6 | 7 | 8 | 9 | 10 |
| --- | --- | --- | --- | --- | --- | --- | --- | --- | --- | --- |
| $n$ | 9 | 139 | 9 | 13 | 0 | 0 | 0 | 0 | 0 | 0 |
| % | 5.3 | 81.8 | 5.3 | 7.6 | 0.0 | 0.0 | 0.0 | 0.0 | 0.0 | 0.0 |

All three exact-agreement MST 5 images were allocated as archetypes. The evaluation set therefore covers only MST 1–4.

The 81.8% concentration on MST 2 is the principal driver of the constant-baseline result reported in Section 4.2.

## 4 Three Pipelines and the Main Result

This section specifies the three pixel-based MST annotation pipelines we evaluate, the shared front-end and calibration step, and the main result: all three pipelines produce real ordinal signal (bootstrap confidence intervals exclude zero) but fall short of sufficient performance on the full evaluation set, where the 82% MST-2 skew gives a constant-mode baseline an insurmountable accuracy floor. A class-balanced bootstrap analysis isolates the algorithmic signal from the dataset bias. We then diagnose the performance ceiling by inspecting the per-class prediction distribution and the extracted Lab clouds.

### 4.1 Three families, one shared front-end

Pixel-based prior work (Section 2.5) covers two families: *(a) palette matching*, in which a representative image colour is extracted by clustering and matched to the nearest reference anchor in a perceptual colour space [15]; and *(c) 1D colorimetric scalars*, in which the colour is projected to the Individual Typology Angle (ITA) and matched in 1D [3, 9]. We add *(b) robust statistics* (a trimmed-mean estimator over the skin pixels, with matching restricted to the chromatic axes that dermoscopy is least likely to corrupt) as a third family to test an intermediate extraction hypothesis not previously applied to this modality. Supervised classifiers (Section 2.5) are excluded: they require in-domain labelled training data that does not exist for dermoscopy MST annotation. We implement one representative pipeline per family on a shared front-end and evaluate all three under the same conditions.

All three pipelines share a three-stage front-end (segmentation, perilesional ring, artifact mask), the retained pixels are then converted to CIE Lab as the input to extraction, and differ only at stages 4 and 5 (colour extraction and anchor matching); the form of the stage 3 artifact mask differs by method as described below. The three end-to-end pipelines are shown in Figures 4 to 6, one per method, alongside the corresponding paragraph and pseudocode below. We treat the three pipelines as representatives rather than optimised proposals: the feasibility finding depends on what every member of these families inherits from the modality, not on a particular hyperparameter choice.

**Figure 4:**
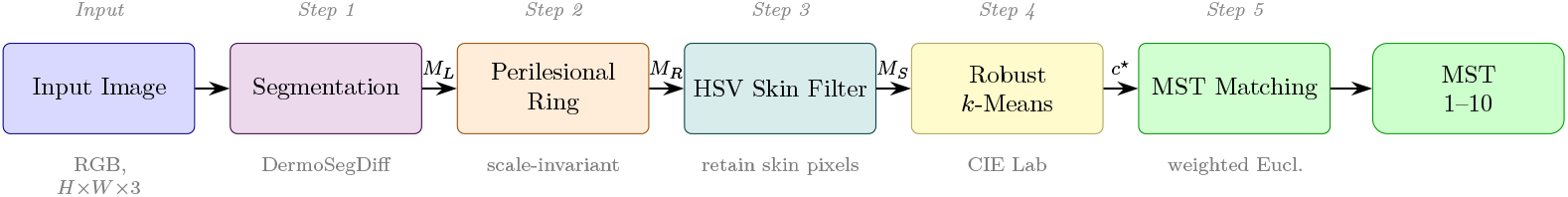
Palette pipeline. The input dermoscopy image enters at left. (1) Lesion segmentation produces *M*_*L*_. (2) A scale-invariant perilesional ring *M*_*R*_ is dilated from *M*_*L*_. (3) An HSV skin filter intersects with *M*_*R*_ to give the skin mask *M*_*S*_. (4) Pixels under *M*_*S*_ are converted to CIE Lab and clustered by *k*-means; the largest plausible cluster yields a dominant colour *c*^⋆^. (5) The MST label is the nearest anchor to *c*^⋆^ under weighted Euclidean distance (*L*\*-weight 3).

One sharing relationship is worth noting explicitly. TrimLab and ITA use *identical* colour extraction (trimmed-mean over the same 25% tail, same pixel set): they differ only at the matching stage (2D Euclidean on (*L*\*, *b*\*) vs. 1D ITA interpolation). They therefore probe distinct matching hypotheses on a shared extraction, not two independent families from extraction onward. Palette is more distinct: it uses *k*-means cluster centroids rather than a trimmed mean, and its results vary across *k* values (we sweep *k* ∈ {1, …, 5}).

#### Shared front-end (stages 1–3)

Stage 1 segments the lesion with DermoSegDiff [2]. Stage 2 dilates the lesion mask by an inner buffer of 0.15 *d* and an outer extent of 0.40 *d*, where 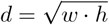 pixels from the lesion bounding box (measured on the 128 × 128 resized image), and takes the difference to produce a scale-invariant perilesional ring of width 0.25 *d*. Stage 3 filters the ring for artifacts using a method-specific mask, because Palette and TrimLab/ITA handle artifact rejection at different stages. For Palette, this is an HSV skin filter (*H* ∈ [0, 25], *S* ∈ [40, 255], *V* ∈ [50, 255]) that retains only skin-coloured pixels, falling back to the unfiltered ring if fewer than *τ*_skin_ pixels survive. For TrimLab and ITA, which reject artefacts via *L*\* trimming at extraction, stage 3 applies only a vignette mask (grayscale threshold *V >* 15) to exclude near-black image borders. The retained pixels are then converted to CIE Lab as the input to extraction. Algorithm 1 gives the pseudocode (with ArtifactMask standing for the method-specific stage 3 filter).

##### Algorithm 1

ExtractSkinPixels (shared front-end).

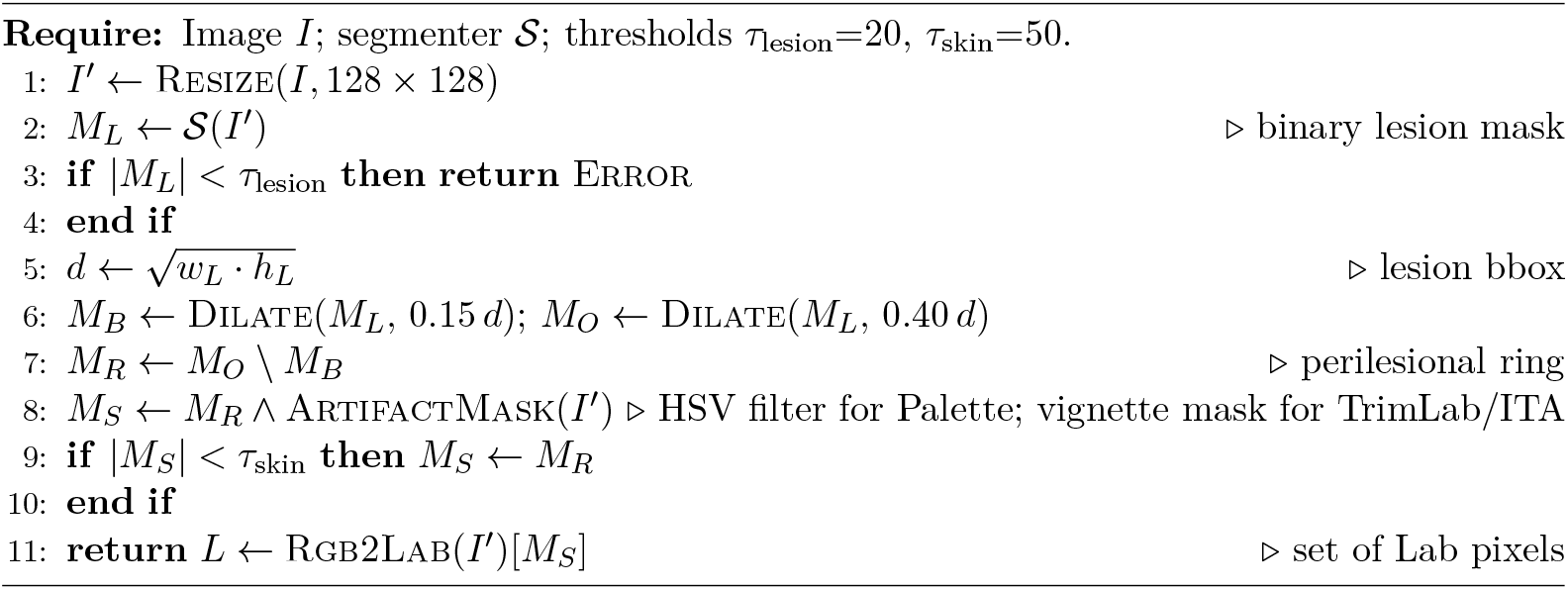

#### Palette: *k*-means clustering with weighted Euclidean distance

Dermoscopy images vary substantially in illumination across cameras and immersion fluid, so *L*\* values in the perilesional ring are normalised by their 90th-percentile before clustering, rescaling to a fixed canonical reference (*L*\*=92). *k*-means is then run on the normalised skin pixels; we report *k*=2 as the representative cluster count, the value maximising QWK on the evaluation set across the sweep *k* ∈ {1, …, 5}. The largest cluster whose centroid passes a skin-plausibility gate (*L*\* ∈ [40, 95], *a*\* > − 5, *b*\* > − 5) is taken as the dominant colour *c*^⋆^; the gate excludes clusters anchored on hair, shadow, or gel. If no cluster passes, the lightest cluster serves as fallback, since shadows and hair are always darker than perilesional skin in dermoscopy. Matching uses a weighted Euclidean distance that up-weights *L*\* by a factor of 3, because MST is fundamentally a lightness scale and dermoscopy shifts *a*\* and *b*\* systematically through polarisation and gel optics:

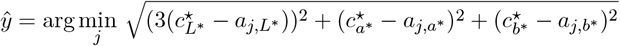

where *a*_*j*_ is the *j*-th MST anchor in Lab. This follows the palette-matching approach of Thong et al. [15] with a weighted Euclidean distance in place of CIEDE2000. Figure 4 shows the end-to-end pipeline; Algorithm 2 gives the pseudocode.

##### Algorithm 2

Palette (*k*-means + weighted Euclidean). Run for *K* ∈ {1, …, 5}; *K*=2 is the representative result.

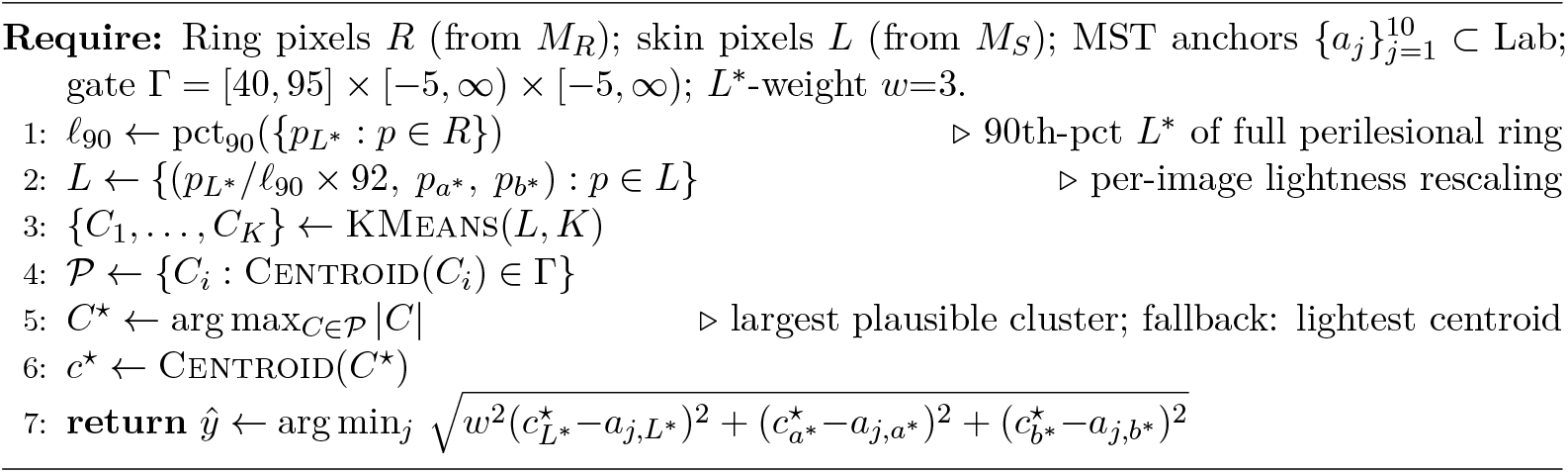

#### TrimLab: trimmed-mean extraction with 2D Lab matching

A representative skin colour *c*^⋆^ is extracted by sorting the perilesional pixels by *L*\* and taking the component-wise mean of the middle 50% in Lab. This trimmed estimator explicitly discards the bottom 25% (typically hair and shadow) and the top 25% (typically glare and immersion-fluid highlights). Matching is then performed using a 2D Euclidean distance on (*L*\*, *b*\*), deliberately ignoring the *a*\* axis. This tests the hypothesis that perilesional erythema systematically contaminates *a*\* and misleads the matcher. As a pipeline, TrimLab applies illumination-aware robust statistics to recover the underlying skin tone. Both extraction and matching are deterministic (no random sampling). Figure 5 shows the end-to-end pipeline; Algorithm 3 gives the pseudocode.

**Figure 5:**
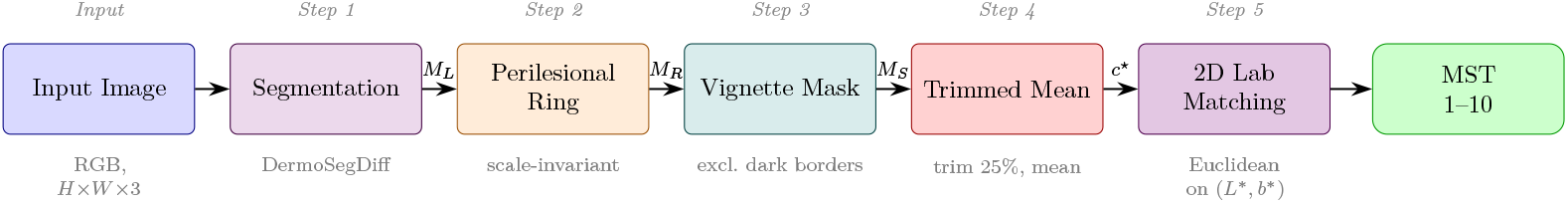
TrimLab pipeline. Steps 1 and 2 follow Figure 4; step 3 applies a vignette mask (grayscale threshold *V >*15) in place of the HSV skin filter. (4) Skin pixels are sorted by *L*\* and the middle 50% are component-wise averaged in Lab to produce *c*^⋆^, replacing *k*-means with a robust statistic. (5) The MST label is the nearest anchor under 2D Euclidean distance on (*L*\*, *b*\*), ignoring the *a*\* axis to suppress perilesional-erythema confounding.

##### Algorithm 3

TrimLab (trimmed-mean + 2D Lab).

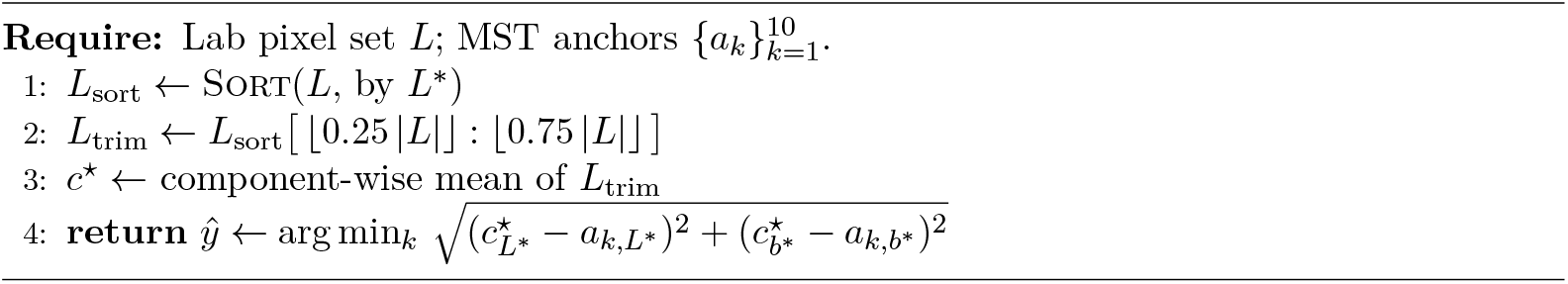

#### ITA: trimmed-mean extraction with ITA scalar projection

The colour extraction is identical to TrimLab: the same trimmed-mean *c*^⋆^ is computed over the middle 50% of the *L*\* distribution. This colour is then projected to the Individual Typology Angle, ITA =arctan((*L*\* − 50)/*b*\*)·180/π, with *b*\* clamped to ≥ 1 to prevent numerical instability on near-achromatic skin. Matching is performed by passing this scalar through a 1D linear interpolator built over the ITA values of the canonical anchors, with the output rounded to the nearest integer in {1, …, 10}. Because ITA is an established, objective colorimetric index for skin tone in dermatology [3, 9], this pipeline tests whether collapsing the colour space into a single lightness-to-yellowness ratio mitigates dermoscopy’s chromatic distortion. Both extraction and matching are deterministic (no random sampling). Figure 6 shows the end-to-end pipeline; Algorithm 4 gives the pseudocode.

**Figure 6:**
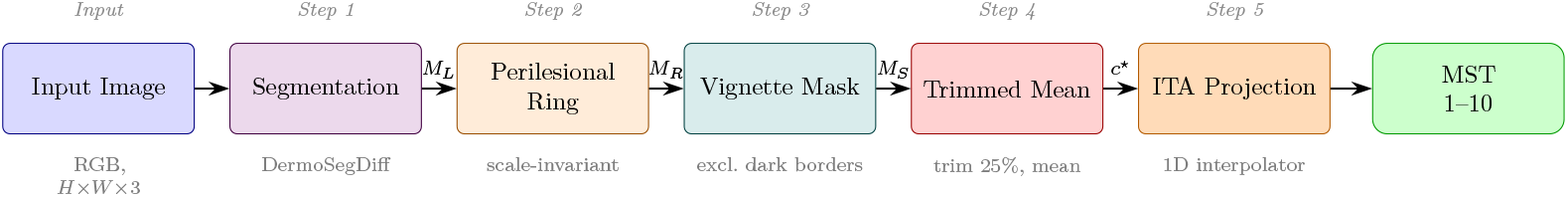
ITA pipeline. Steps 1–4 are identical to Figure 5 (the same trimmed-mean extraction). (5) The dominant colour *c*^⋆^ is projected to the Individual Typology Angle ITA = arctan((*L*\*− 50)*/b*\*)·180*/π*, then mapped to an MST label via a 1D linear interpolator *ϕ* over the canonical-anchor ITAs.

##### Algorithm 4

ITA (trimmed-mean + ITA projection).

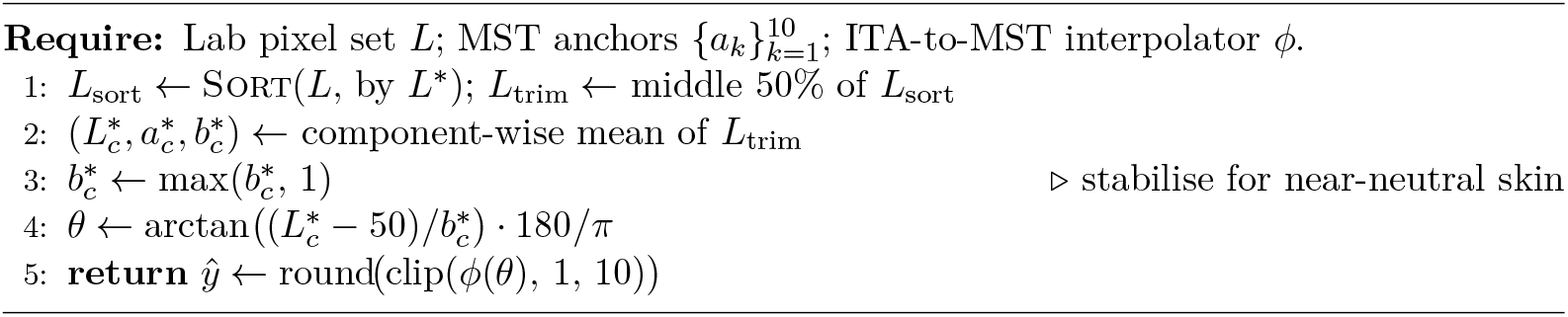

#### Calibration (shared)

Each method is evaluated in two variants: *uncalibrated*, in which the matching anchors are the canonical sRGB MST values [12] converted to Lab, and *calibrated*, in which the per-class anchors are replaced by the component-wise median of *c*^⋆^ on that class’s archetype set 𝒜_*k*_:

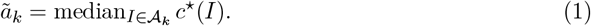

Here *c*^⋆^(*I*) is the Lab vector produced by each method’s colour-extraction step. For Palette and TrimLab, ã_*k*_ is used directly as a Lab anchor in the matching step. For ITA, calibration is performed in the projection space: *c*^⋆^(*I*) is first projected to a scalar *θ*(*I*) = ITA(*c*^⋆^(*I*)), and the calibrated anchor for class *k* is 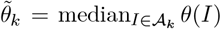. The 1D interpolator *ϕ* is rebuilt over 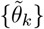before inference. Palette and TrimLab calibration operates in Lab; ITA calibration operates in the same scalar space used for matching. The three methods handle classes without archetypes (MST 6–10 in our split) differently. Retaining canonical anchors for un-archetyped classes alongside calibrated anchors for the rest would mix two distinct colour distributions in a single matching set and systematically bias matching toward the uncalibrated classes; all three methods therefore avoid that mixture. Palette drops MST 6–10 from the matching set entirely, restricting predictions to the calibrated 1–5 range. TrimLab and ITA instead linearly extrapolate the calibrated MST 1–5 anchors to fill MST 6–10 (component-wise on (*L*\*, *a*\*, *b*\*) for TrimLab; on the ITA scalar for ITA). Extrapolation extends the calibrated manifold rather than excluding it, keeping every anchor in a single calibrated colour distribution while allowing predictions to reach beyond MST 5 when the extracted colour trends darker than any archetype. In practice the extrapolated classes are rarely selected on this evaluation set: TrimLab predicts no eval image at MST ≥6, and ITA predicts one. Table 3 summarises the hyperparameters; full configurations are released alongside the code.

**Table 3:** Pipeline hyperparameters. Stages 1–3 and the archetype split are shared. Stages 4 and 5 differ by method.

| Symbol | Value | Meaning |
| --- | --- | --- |
| *Shared (stages 1–3)* |  |  |
| Working resolution | $128 \times 128$ | Pipeline operating size |
| $\tau_{\text{lesion}}$ | 20 px | Min lesion mask size |
| $\tau_{\text{skin}}$ | 50 px | Min skin mask size (100 px for Palette) |
| Buffer ratio | 0.15 | Inner buffer, fraction of $d$ |
| Ring ratio | 0.25 | Ring width, fraction of $d$ |
| Segmenter | DermoSegDiff | ISIC 2018 weights |
| *Palette (k-means + weighted Euclidean)* |  |  |
| HSV hue | [0, 25] | Skin-filter hue range |
| HSV saturation | [40, 255] | Skin-filter saturation range |
| HSV value | [50, 255] | Skin-filter value range |
| $k$ | 2 | $k$-means cluster count (swept $k \in \{1, \dots, 5\}$) |
| $L^*$ normalisation | 90th-pct $\rightarrow L^*=92$ | Per-image lightness rescaling |
| $L^*$ gate | [40, 95] | Plausible skin lightness |
| $a^*$ / $b^*$ gate | $> -5$ | Reject blue/green clusters |
| Matching distance | Weighted Euclidean | $L^*$-weight = 3 |
| *TrimLab (trimmed-mean + 2D Lab)* |  |  |
| Vignette threshold | $V > 15$ | Exclude near-black borders |
| Trim fraction | 25% | Each tail of $L^*$ distribution |
| Matching distance | 2D Euclidean | On $(L^*, b^*)$ |
| *ITA (trimmed-mean + ITA projection)* |  |  |
| Vignette threshold | $V > 15$ | Exclude near-black borders |
| Trim fraction | 25% | Each tail of $L^*$ distribution |
| $b^*$ clamp | $\geq 1.0$ | For ITA stability |
| Matching | 1D interpolator | Anchor ITA → MST |

### 4.2 Main result: real signal, insufficient performance

We compare the six (method × variant) pipeline configurations against two trivial baselines: a constant prediction of MST 2 (the mode of the eval set) and a random predictor that draws each prediction independently from the eval-set class distribution (MST 2 with probability 82%, MST 4 with 8%, MST 1 and 3 each 5%). For the random predictor we report the analytical expectation under this sampling distribution (equivalently, the large-sample mean over independent draws); single-seed runs vary around these values. Table 4 reports the four ordinal-classification metrics on the 170-image evaluation set; Figure 7 visualises them.

**Table 4:** Pipeline performance on the 170-image evaluation set. The best (non-ceiling) value in each column is bolded. All three methods report a single deterministic run; Palette uses *k*=2 as the representative cluster count. The constant “Always MST 2” baseline dominates on Exact, Within-1,and MAE because 82% of images are MST 2; QWK is identically zero by construction for any constant predictor. Bootstrap 95% confidence intervals for QWK are reported in Table 5. Palette metrics are computed on 169 images; one eval-set image (ISIC_0026403) has fewer than *τ*_skin_=100 usable perilesional pixels (both the HSV-filtered mask and the raw ring fall below the threshold) and is excluded.

| Method | Exact (%) | $\leq 1$ shade (%) | MAE | QWK |
| --- | --- | --- | --- | --- |
| **Always MST 2** | **81.8** | **92.4** | **0.26** | 0.000 |
| Random (freq. weighted) | 68.0 | 86.1 | 0.47 | 0.000 |
| Palette uncal | 53.3 | 62.7 | 1.04 | −0.010 |
| TrimLab uncal | 59.4 | 75.9 | 0.93 | 0.163 |
| ITA uncal | 8.2 | 87.1 | 1.11 | 0.104 |
| Palette cal | 19.5 | 68.6 | 1.16 | 0.191 |
| TrimLab cal | 36.5 | 78.2 | 0.87 | **0.211** |
| ITA cal | 40.6 | 85.9 | 0.77 | 0.197 |
| Human IRR (eval set) | 84.1 | 97.1 | 0.19 | 0.699 |

**Figure 7:**
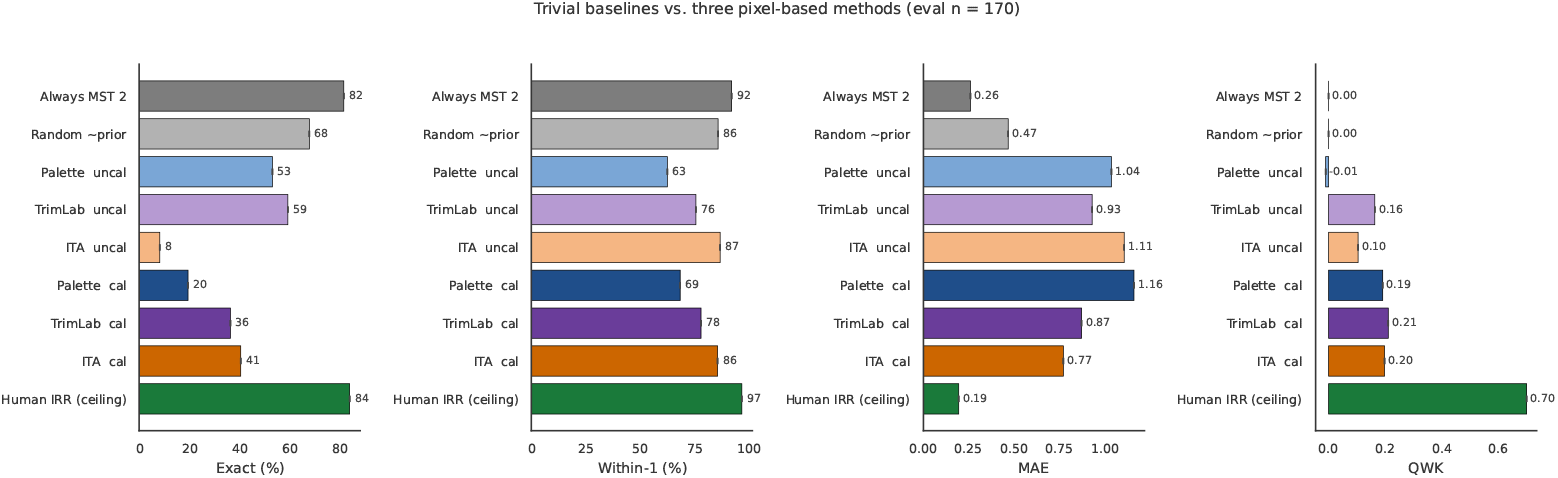
Trivial baselines vs. three pixel-based methods. Across exact match, within-one-shade, and MAE, every calibrated pipeline (dark blue, dark purple, dark orange) is dominated by the “always predict MST 2” constant baseline. Calibrated pipelines win only QWK, where constants are pegged at *κ* = 0 by construction. Palette uses a representative *k*=2. The human inter-rater ceiling (green) is computed on the same eval set.

Three conclusions follow from Table 4. First, **all three pipelines produce real ordinal signal**: bootstrap 95% confidence intervals on QWK are [0.09, 0.34] (TrimLab), [0.07, 0.33] (ITA), and [0.07, 0.30] (Palette), all excluding zero. Second, algorithmic effort across the three families produces genuine gains: calibrated QWK rises from 0.19 (Palette) to 0.21 (TrimLab), and calibrated MAE drops from 1.16 (Palette) to 0.77 (ITA), a 34% reduction. Third, none of those gains are sufficient: a constant “always predict MST 2” classifier beats every calibrated pipeline by at least 41 percentage points on exact match, at least 6 points on within-one-shade, and at least 3× on MAE, because 82% of the eval set is MST 2 and the signal strength does not overcome that floor. The three calibrated QWKs land at 0.19–0.21, between one-quarter and one-third of the human ceiling (0.699). We report ITA as the strongest single configuration because it dominates TrimLab on exact match (41% vs. 36%), within-one-shade (86% vs. 78%),and MAE (0.77 vs. 0.87); TrimLab’s edge in QWK (0.211 vs. 0.197) is within rounding.

The uncalibrated variants illustrate distinct failure modes. Palette has negligible QWK (−0.01), producing predictions ordinally uncorrelated with the consensus. TrimLab reaches QWK 0.16 by concentrating on MST 2, inflating exact match (59%) without genuine discrimination. ITA collapses in the opposite direction, predicting MST 1 on 83% of images and mechanically achieving 87% within-one-shade while contributing no ordinal signal. None improves substantively over a constant predictor.

#### Bootstrap confidence intervals and class-balanced analysis

Table 5 reports 2 000-iteration bootstrap results (each iteration draws 170 images at random, allowing repeats) for the three calibrated methods on two sampling regimes. On the full eval set (left half), all three QWK confidence intervals exclude zero. The methods are not statistically distinguishable from each other: all three CIs substantially overlap. On the class-balanced regime (right half), we stratify by resampling nine images per class from the MST 1–4 evaluation-set images (drawing from the 170-image evaluation set allowing repetition; *n*=36 per iteration), which holds the class distribution constant and isolates the algorithmic signal from the evaluation-set skew.Under this regime, all three methods beat the trivial baseline with CIs excluding zero (TrimLab 0.38 [0.08, 0.62], ITA 0.43 [0.14, 0.66], Palette 0.40 [0.11, 0.64] vs. 0.00). Exact match improves to 33–42% vs. 25% for the trivial baseline. This confirms that all three methods produce genuine ordinal signal, not an artifact of the MST-2 skew. The gap between the full-eval and balanced-eval pictures is therefore explained almost entirely by dataset representativeness, not by the algorithm family.

**Table 5:**
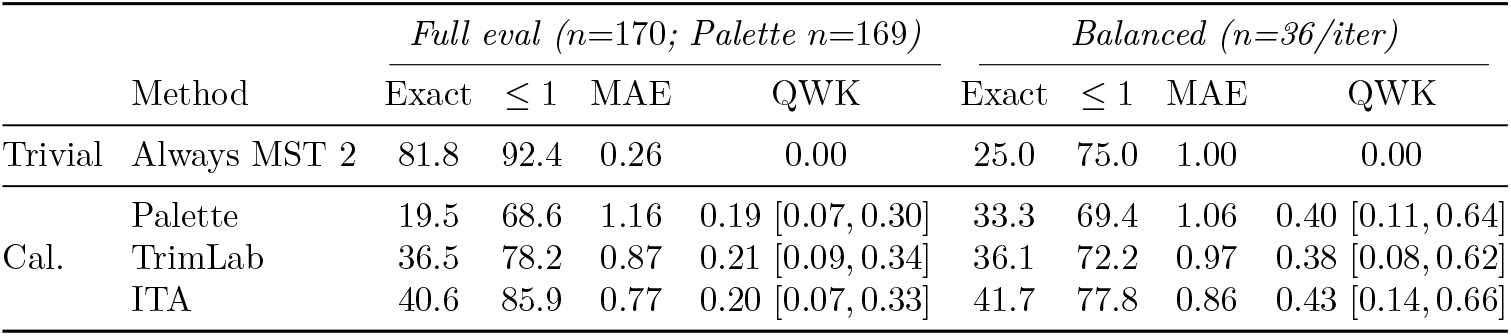
Bootstrap analysis (2 000 iterations). Left: resampling with replacement from the full eval set (*n*=170; Palette *n*=169, see Table 4), 95% CIs in brackets. Right: stratified resampling of 9 images per class from MST 1–4 (*n* = 36 per iteration), removing the 82% MST-2 skew. All three calibrated QWK CIs exclude zero on both regimes. On the balanced regime all three methods beat the trivial “Always MST 2” baseline with CIs excluding zero. The three methods are not statistically distinguishable from each other on either regime.

### 4.3 Diagnosis: where the performance ceiling lies

Two analyses identify the causes of the performance ceiling shared across all three methods.

#### The dominant class limits absolute-accuracy metrics

Figure 8 shows the calibrated prediction distribution for true MST 2 (which accounts for 82% of the eval set) under each method. Palette concentrates on MST 1 and MST 4 (47%/15%/6%/30%); TrimLab starts to peak on the correct class (37% on MST 2) but still splits substantial mass to MST 1 (29%) and MST 4 (19%); ITA peaks higher (41% on MST 2) but is otherwise comparable. The algorithmic progression is real (ITA places 41% of mass on the correct class vs. 15% for Palette), but when 82% of images are MST 2, the constant baseline places 100% of mass there by construction. No level of ordinal signal at the current magnitude can overcome this floor on a distribution this skewed.

**Figure 8:**
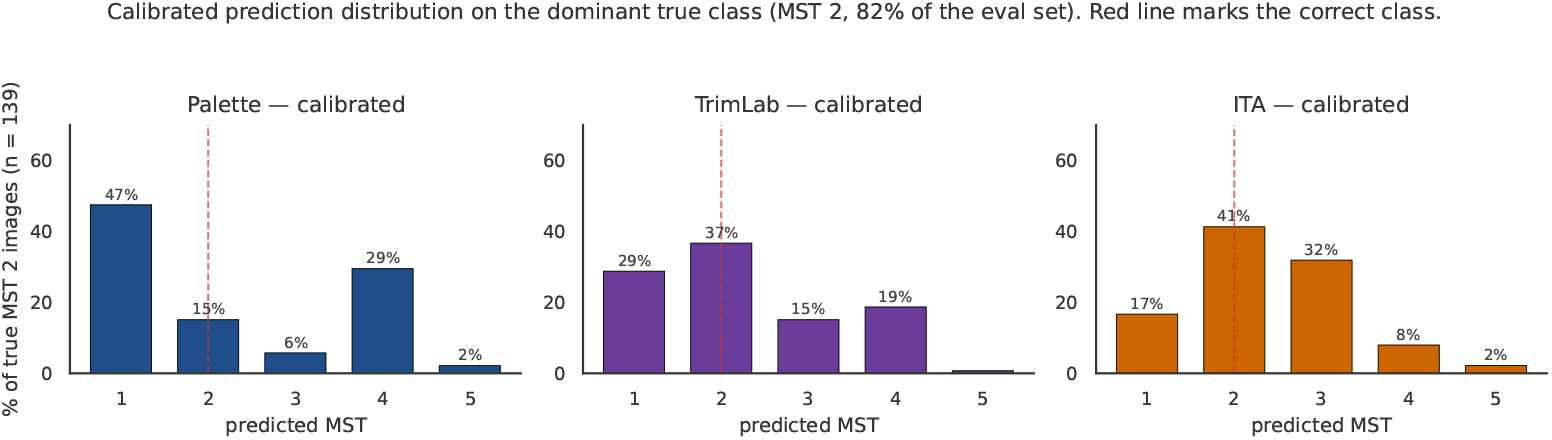
Calibrated prediction distribution on the dominant true class (MST 2). Each panel shows, for true-MST-2 images (n= 139, 82% of the eval set), what fraction the corresponding calibrated pipeline predicts at each MST label. Red dashed line marks the correct class. Algorithmic improvement across Palette, TrimLab, ITA raises the mass at MST 2 from 15% to 37% to 41%, but none of the three concentrates on the truth at the rate that a constant predictor would.

#### Chromatic overlap limits discrimination at the light end

Figure 9 plots the extracted Lab clouds (one circle per eval-set image, coloured by consensus class) for Palette (*k*-means cluster centroid) and ITA (trimmed-mean extraction; TrimLab uses the same extracted colour as ITA and is omitted for legibility). The canonical anchors for MST 1 and MST 2 are only 2.7 Lab units apart; MST 1 and MST 3 are 9.0 Lab units apart, a gap dominated by Δ*b*\* = 8.8.Under *both* extraction strategies, the consensus-class clouds for MST 1, MST 2, and MST 3 overlap heavily on the chromatic axes: within-class standard deviations are comparable to or larger than the inter-anchor margin. The *k*-means extraction (Palette, left) sits closer to the canonical anchors but is highly dispersed, the trimmed-mean extraction (ITA, right) is tighter but systematically shifted to lower *L*\* relative to the lighter anchors. The chromatic signal that distinguishes MST 1–3 has been compressed by the imaging modality, limiting discrimination at the light end of the scale; this is a property of the input that is shared across all three matching strategies. Note that the class-balanced bootstrap (Table 5) confirms that signal does survive above MST 2, and that the overlap is concentrated in the MST 1–3 range where the inter-anchor margin is smallest.

**Figure 9:**
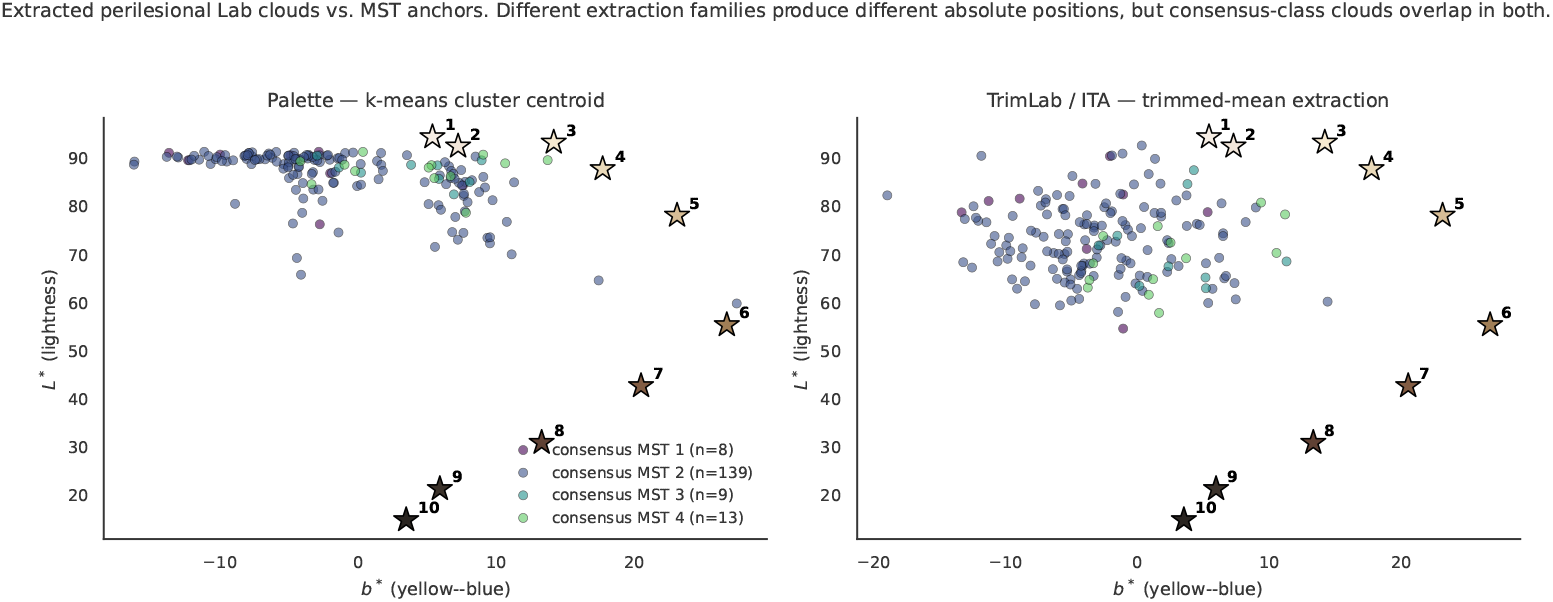
Extracted perilesional skin colour vs. MST reference colours, plotted in the *b*\* (yellow–blue) vs. *L*\* **(lightness) plane**. Each circle is one evaluation-set image, positioned by the skin colour extracted from its perilesional ring and coloured by the human consensus MST label. Stars mark the ten canonical MST reference colours (numbered). The algorithm assigns each image the label of its nearest star, so when circles of different colours overlap the algorithm cannot reliably distinguish those classes. Left: Palette extracts the *k*-means cluster centroid. Right: TrimLab and ITA both extract the trimmed-mean colour (shown together). Under both extraction strategies, the circles for MST 1, 2, and 3 overlap heavily: dermoscopy’s cross-polarised illumination compresses the *b*\* range, collapsing the separation between lighter skin tones that exists in the reference colours.

### 4.4 Confusion matrices

Figure 10 shows the calibrated confusion matrices for the three methods, restricted to MST 1–5 (the only classes that appear in the eval set). The matrices visualise the algorithmic progression we report quantitatively in Table 4: Palette disperses true-MST-2 predictions across MST 1, MST 2, MST 3, and MST 4 with comparable mass; TrimLab begins to concentrate on MST 2 (51 of 139 true-MST-2 images correctly classified, vs. 21 in Palette); ITA concentrates further (57 of 139). The progression demonstrates that the engineering improvements between methods are visible on the confusion structure as well as on the aggregate metrics. None of the three reaches the > 80% correct-on-MST-2 mass that the trivial baseline does mechanically.

**Figure 10:**
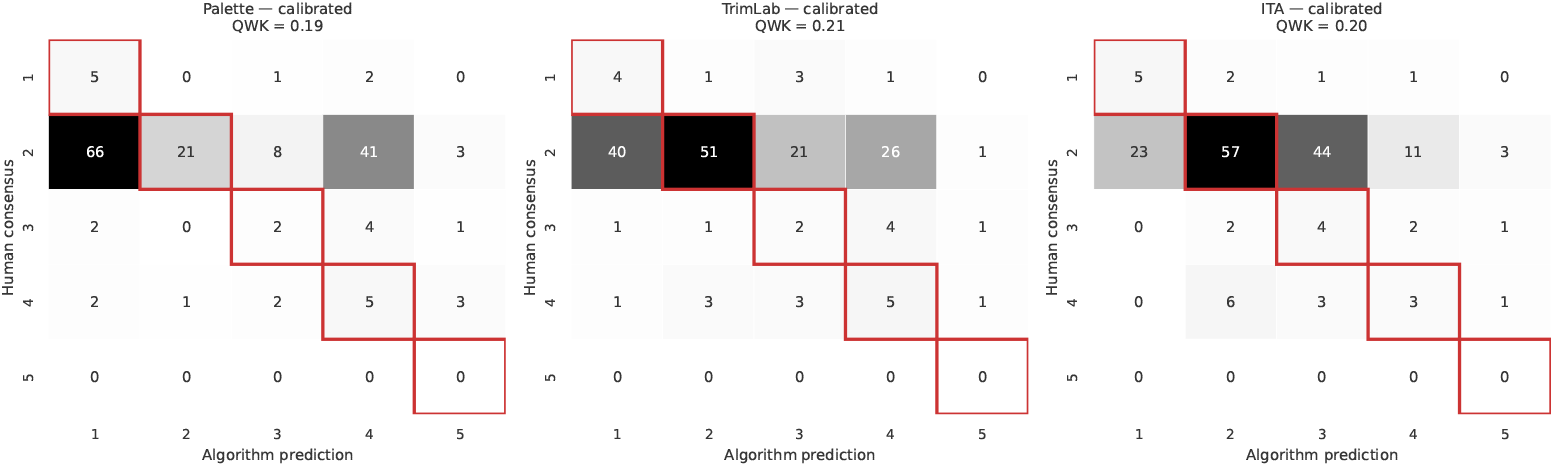
Calibrated confusion matrices for the three methods on the 170-image eval set. Rows: human consensus labels; columns: pipeline predictions; values show Palette (*k*=2), TrimLab, and ITA. Red outlines mark the diagonal (correct matches). The progression Palette → TrimLab → ITA increases the correct-on-MST-2 count from 21 to 51 to 57, but none reaches > 80% of the 139 true-MST-2 images.

### 4.5 What we ruled out

We considered four alternative explanations and tested each.

#### Segmentation quality

If the perilesional ring sometimes falls outside the skin region, extracted Lab would be biased toward non-skin colours. We visually inspected perilesional masks for a convenience sample of misclassified images; in all examined cases the ring sat on visible skin. This is not a systematic audit: we cannot rule out occasional mask failures on individual images, but we expect these to be rare given DermoSegDiff’s Dice score of 0.90 on the ISIC 2018 segmentation challenge [2]. Segmentation quality is unlikely to be the primary performance bottleneck.

#### Hyperparameter sensitivity (Palette)

We swept *k* ∈ {1, …, 5}. QWK across the sweep ranges from 0.09 (*k*=3) to 0.19 (*k*=2); we report *k*=2 as the representative value. Even at *k*=2, calibrated Palette (QWK = 0.19) remains dominated by the constant baseline on absolute-accuracy metrics, so the performance ceiling is not a hyperparameter-search artifact.

#### Distance metric (Palette)

We compared our weighted Euclidean distance (*L*\*-weight 3) against CIEDE2000. Performance was comparable, confirming that the choice of distance metric is not the limiting factor: the chromatic signal needed to distinguish MST 1–3 has been removed before the matcher sees the input.

#### Algorithmic family

The three methods (Palette cluster matching, TrimLab robust statistics with chromatic axes, ITA 1D colorimetric scalars) reflect distinct hypotheses about which family of pixel-based methods would absorb the dermoscopy distortion. None reaches sufficient performance, and bootstrap confidence intervals confirm the three are statistically indistinguishable from each other (Table 5). The performance ceiling is therefore shared across this family of methods, and is not specific to the choice of extractor, distance metric, or scalar projection.

## 5 Discussion

This section scopes the feasibility finding, identifies modalities for which we expect pixel-based MST estimation to remain viable, and discusses caveats to the benchmark design.

### 5.1 What the feasibility study shows, and what it does not

The result we report has two components. First, Lab-based pixel features carry real MST signal on dermoscopy: bootstrap confidence intervals exclude zero for all three calibrated methods on the full eval set, and a class-balanced bootstrap confirms ITA achieves QWK 0.43 against a trivial baseline of zero (Table 5). Second, the signal is not sufficient for autonomous annotation at the scale and distribution of ISIC 2018: on the full eval set (82% MST 2), all three calibrated methods are dominated by a constant-mode predictor on exact match and MAE. These two components point to different follow-up directions and generalise separately.

#### Generalisation across pipelines, within dermoscopy

The diagnostic in Section 4.3 shows that dermoscopy’s cross-polarized illumination causes the extracted colours for MST 1, 2, and 3 to overlap, making lighter skin tones hard to distinguish regardless of which algorithm is used. The three methods we tested each take a different approach to working around this: palette matching with *k*-means cluster centroids (Palette), robust trimmed-mean extraction with chromatic-axis matching (TrimLab), and and projection to a 1D colorimetric index (ITA). None reaches sufficient performance, and bootstrap CIs confirm they are statistically indistinguishable from each other. We therefore expect this ceiling to apply to any method that compares extracted pixel colours to reference anchors, regardless of the specific distance metric or extractor used. We do *not* claim that supervised approaches, such as a CNN trained end-to-end on dermoscopic MST labels, would also be limited; the question is open and would require substantially more annotated data than the benchmark provides.

#### Generalisation across datasets, within dermoscopy

ISIC 2018 is the largest public dermoscopy dataset and the principal training corpus for downstream skin-lesion classification, so a finding specific to ISIC 2018 dermoscopy already covers most published benchmarks. Other dermoscopy datasets (PH^2^, ISIC 2019/2020, HAM10000) are captured with comparable equipment and inherit the same chromatic-compression property. We expect the performance ceiling to transfer, we have not tested it.

#### Generalisation across modalities

The feasibility finding does *not* extend to clinical photography (non-dermoscopic skin photographs taken under ambient light) or to selfies. Both have richer chromatic dynamic range and have been the primary modality for prior MST and Fitzpatrick estimation work [6, 8, 15]. Pipelines very close to those we evaluate, including ITA,which has a long track record on clinical photography [9], are likely viable on those modalities; this is an empirical claim of prior literature, not something we test here. The most direct follow-up to this study is re-evaluating the same three pipelines on a clinical-photo dataset (e.g., DDI [6], Fitzpatrick 17k [8], or PAD-UFES-20), where the richer chromatic dynamic range should enable substantially better performance and would isolate the dermoscopy-specific compression effect from the algorithmic design.

### 5.2 The high NA rate and label coverage

A 60% NA rate is the data point that most directly constrains practical deployment of any dermoscopy MST estimator. All three pipelines produce a numeric label for every image whose lesion segmentation passes *τ*_lesion_ = 20 pixels; this gate excludes ∼1% of ISIC 2018, not 60%. The other 59% are images where a human, free to zoom and consult the swatch reference, judged the visible perilesional skin too occluded to label, but where the pipelines confidently emit a number. For downstream uses such as fairness audits, where the consumer of the label wants to stratify a training set or report subgroup metrics, this asymmetry creates a critical structural vulnerability: the label looks complete, but it is not verifiable on the majority of images.

We see two acceptable responses to this. First, any pixel-based pipeline applied to dermoscopy should be modified to abstain when the visible perilesional skin is small, hair-occluded, or low-contrast, matching the criteria the human raters used. Designing a calibrated abstention rule is a non-trivial follow-up. Second, the consumer of the labels should treat algorithmic MST predictions on dermoscopy as a noisy proxy with a known failure floor (the constant baseline) and report uncertainty intervals accordingly. Neither response is implemented in this study.

### 5.3 The narrow MST range

The 193-image consensus subset contains no images above MST 5. This is not a flaw of the protocol or the sample size; it is a property of ISIC 2018 that the benchmark exposes. Two consequences follow.

First, the benchmark cannot evaluate a pipeline’s behaviour on darker-skinned images, which is the area where fairness concerns are sharpest. This study says nothing about MST 6–10 performance on dermoscopy, in either direction. A representative evaluation requires a darker-skinned dermoscopy cohort, which to our knowledge does not exist publicly at the scale needed for dual-rater ground truth. Building one is an important community task.

Second, the within-1-shade and exact-match metrics are weakly informative on a heavily skewed eval set. The trivial-baseline analysis in Section 4.2 uses this property to set a high accuracy floor that any pipeline must exceed. On a darker or more uniform cohort, the same baseline would be much weaker, and the absolute pipeline numbers would also change. We report all four metrics, with the constant baseline alongside, precisely so that a reader can re-anchor expectations to a different distribution if a darker cohort becomes available.

### 5.4 The benchmark itself

#### Two raters

The MST-Derm consensus is two-rater. Two raters is the minimum at which inter-rater reliability can be reported, and it is the standard in published MST work [12], but a three-rater consensus would be more robust.

#### Annotator pool

Both raters were non-clinicians drawn from a small pool, and rater demographics including self-reported skin tone were not recorded. MST labelling is a colour-matching task against a fixed palette and does not require dermatological expertise, but raters with different self-reported skin tones may calibrate the swatch reference differently, and a demographically diverse rater pool would strengthen external validity. We flag this as a limitation, future annotation rounds using this protocol should record rater demographics.

#### The pipelines as representatives

We described each of the three pipelines as “representative” of its methodological family rather than “optimal” for it. The performance ceiling depends on what every member of the pixel-distance family inherits from the modality (compressed *b*\* for MST 1–3), not on a particular extractor or distance choice. A reader who suspects a specific alternative (a different segmenter, a different *k*, a different trim fraction, a non-linear ITA-to-MST mapping) can run it on the released eval set and compare directly. As noted in Section 4.1, TrimLab and ITA share the same colour-extraction step and differ only at matching, so a reader specifically interested in separating extraction from matching effects can compare their results as a matched pair. We have made the deterministic and seed-specific prediction CSVs for all three methods public for exactly this comparison.

## 6 Conclusion

We released MST-Derm, a dual-rater Monk Skin Tone benchmark on 500 ISIC 2018 dermoscopy images, with a 193-image consensus subset (quadratic-weighted Cohen’s *κ* = 0.82), an explicit *unrateable* option used by raters on 60% of the underlying sample, and a 170-image evaluation set held out from a calibration archetype split. We evaluated three pixel-based pipelines on this benchmark, one per principal methodological family in the prior literature, sharing a front-end and an archetype-based calibration step: *k*-means clustering with weighted-Euclidean anchor matching (Palette), trimmed-mean extraction with 2D Lab matching on (*L*\*, *b*\*) (TrimLab),and trimmed-mean extraction with Individual Typology Angle interpolation (ITA).

The feasibility study yields two findings. First, **Lab-based pixel features carry real MST signal on dermoscopy**: bootstrap confidence intervals confirm that all three calibrated methods produce ordinal signal statistically above zero (QWK 95% CIs: Palette [0.07, 0.30], TrimLab [0.09, 0.34], ITA [0.07, 0.33]), and a class-balanced bootstrap shows ITA achieves QWK 0.43 against a trivial baseline of zero. Second, **current performance is insufficient for autonomous annotation** on ISIC 2018: every calibrated pipeline is dominated by a constant “always predict MST 2” baseline on exact match (20–41% vs. 82%) and MAE (0.77–1.16 vs. 0.26), because the 82% MST-2 skew gives the constant predictor an accuracy floor the methods cannot overcome. The three methods are not statistically distinguishable from each other: method choice within the pixel-based family does not matter at this scale.

We trace the performance ceiling to two causes. Cross-polarized dermoscopy illumination compresses the chromatic axes (*a*\*, *b*\*) below the inter-anchor margin separating MST 1–3, limiting discrimination at the light end. And ISIC’s light-skin bias, together with NA-driven attrition on hair-dense and darker crops, leaves the evaluation set with 82% MST-2 mass and no images above MST 5. This skew is the primary driver of the gap between the full-eval picture and the balanced-bootstrap picture. The class-balanced analysis establishes that the bottleneck is dataset representativeness, not the algorithm family.

The most direct follow-up is the same three pipelines on clinical photography, where the richer chromatic dynamic range should yield substantially better performance and would isolate the dermoscopy-specific compression. The benchmark, the annotation protocol, the calibration–evaluation split, and all prediction runs are released at https://github.com/Amixdu/dermoscopy-mst. By providing a human-validated ground truth and diagnosing current optical failure modes, we hope this work serves as a foundation for developing robust skin-tone estimators, which are urgently needed to audit and improve fairness in clinical AI.

## Data Availability

The original dermoscopy images used in this study are publicly available from the International Skin Imaging Collaboration (ISIC) 2018 archive. The newly generated MST-Derm benchmark annotations, the evaluation splits, and all algorithmic prediction outputs are publicly available in the project's GitHub repository.

https://github.com/Amixdu/dermoscopy-mst

https://challenge2018.isic-archive.com/

## Acknowledgements

The authors thank Anh Quan Tran for serving as the second independent annotator in the MST-Derm annotation study. This research received no specific grant funding. During the preparation of this work, the authors used Artificial Intelligence to improve the language, readability, and formatting of the manuscript. After using this tool, the authors thoroughly reviewed and edited the content, and take full responsibility for the final publication.

## A Annotation Protocol

This appendix reproduces the written protocol given to both raters verbatim, minus the operational setup instructions (image folder location, CSV field names) that are irrelevant to a reader evaluating the benchmark. The full guide, including worked examples with perilesional-ring overlays, is released at https://github.com/Amixdu/dermoscopy-mst.

### Step 1: Identify the area of interest

First, identify the edge of the main lesion. Mentally create a small buffer (a few millimeters) around the entire border. **Do not assess the skin inside this buffer**. The target for assessment is the healthy-looking skin just outside this buffer zone.

### Step 2: Reject artifacts

Within the area of interest, actively ignore the following:

- The lesion itself.
- Any hair shafts or follicles.
- Surgical ink markings (usually purple, blue, or black).
- Strong shadows or bright, specular reflections.
- Areas of obvious inflammation or redness.

The goal is to assess the healthy, underlying skin tone.

#### Step 3: Compare and decide

Focus on the largest, most uniform patch of healthy, unaffected skin within the area of interest. Zoom in to this area and compare it side-by-side with the MST reference chart. Choose the single MST label (1–10) that is the closest match.

#### Tie-breaker rule

If the colour falls exactly between two MST patches, select the one that better represents the larger portion of the healthy skin in the area of interest.

### Step 4: Unrateable images (NA)

It is crucial **not to guess**. If a confident judgment cannot be made according to the protocol, the image must be marked as unrateable (`NA`).

Use NA if the *entire* area of interest is unusable, for any of the following reasons:

- **Widespread inflammation**. The entire area is uniformly red or otherwise discoloured, with no discernible patch of healthy baseline skin tone visible.
- **Massive obstruction**. The entire area is completely obscured by a large shadow, surgical ink, or other artifact.
- **Poor image quality**. The image is too blurry or out of focus to make a judgment.

